# Routine Data for Workforce Equality Monitoring: Ethnic Inequalities in Recruitment and Workforce Representation in Nursing and Midwifery

**DOI:** 10.64898/2026.03.31.26349776

**Authors:** Anuka Boldbaatar, Sanna Strahle, Azwa Shamsuddin, David Henderson

## Abstract

**Aim:** To examine ethnic inequalities in recruitment outcomes and workforce representation across pay bands among nursing and midwifery staff, and to assess whether routinely collected administrative data can generate reproducible indicators for workforce equality monitoring.

**Design:** Retrospective observational study.

**Methods:** We analyzed routinely collected administrative data from one NHS Board in Scotland. This included annual staff-in-post data for 2021/22 to 2024/25 and pooled recruitment data on interviewed candidates and conditional job offers for 2021/22 to 2023/24. Ethnicity was grouped as White and non-White. Analyses focused on Bands 5, 6 and 7. Recruitment outcomes were assessed using relative risks for receipt of a conditional job offer among interviewed candidates, comparing White and non-White applicants. Workforce representation across pay bands was assessed using representation quotients. Analyses were descriptive and unadjusted.

**Results:** White applicants were more likely than non-White applicants to receive a conditional job offer following interview across all pay bands examined. Inequalities were also evident at Band 5, the usual entry point to registered practice. Workforce composition analyses showed a corresponding gradient in representation, with non-White staff overrepresented in Band 5 and underrepresented in Bands 6 and 7, with little change over the study period.

**Conclusion:** Routinely collected administrative data can generate reproducible indicators of ethnic inequality in recruitment and workforce representation. Embedded within existing workforce systems, such analyses could strengthen workforce equality monitoring, support benchmarking and enhance accountability across healthcare settings.

**Impact:** Utilising routine administrative data for workforce equality monitoring can support policy and practice aimed at improving accountability, retention and workforce sustainability across health systems.

**Reporting Method:** This study followed the Strengthening the Reporting of Observational Studies in Epidemiology (STROBE) reporting guidelines.

**Patient or Public Involvement:** This study did not include patient or public involvement in its design, conduct, or reporting.

## Introduction

Nurses and midwives are central to health service delivery, yet health systems internationally face persistent challenges in sustaining this workforce. Retention has become a major concern in nursing workforce research and policy, with growing evidence of burnout, absence, and turnover risk (1,2). Fair access to recruitment and career progression is an important part of this challenge, because staff experience within organizations helps shape job satisfaction and decisions about whether to remain in the workforce (3). Studies indicate that perceived inequities in career advancement are associated with diminished job satisfaction and increased staff turnover (4,5). Beyond its relationship with turnover, perceived organizational justice has been associated with stronger work engagement and better perceived care quality, suggesting that fairness within organizations may shape not only retention, but also the wider functioning of the workforce (6). More broadly, greater workforce inclusion has been linked to improved organizational performance and patient outcomes (7). Amid growing international mobility and increasing diversity in the health workforce, ensuring equitable career progression is important for both fairness and sustainability. Nevertheless, numerous reports and studies continue to highlight the persistence of ethnic inequalities in recruitment and career progression within the healthcare sector, including underrepresentation of minority ethnic staff in senior roles and persistent barriers to advancement (8,9). These disparities coincide with lower job satisfaction and a higher intention to leave among ethnic minority nurses (10,11). Persistent inequalities cannot be addressed effectively without systematic, comparable data on where disparities occur and how they change over time.

## Background

Summary measures make it possible to compare inequalities across settings and over time (12), while equity indicators derived from administrative data can support organizational benchmarking, quality assurance and action planning (13). However, the extent to which such approaches are embedded in routine workforce monitoring varies across health systems. In England’s National Health Service (NHS), this logic underpins the Workforce Race Equality Standard (WRES) framework which uses standardized workforce race equality indicators to support local, regional and national accountability (14). In Scotland, NHS Boards publish workforce equality information as part of their obligations under the Public Sector Equality Duty (PSED), but there is no equivalent national mechanism for systematic monitoring of workforce ethnic inequalities. Despite repeated calls to strengthen the routine use of workforce ethnicity data (15,16), their use for equality monitoring remains limited by variable data quality, fragmented reporting, and the absence of routine national synthesis. As a result, ethnic inequalities are not consistently analyzed in ways that support benchmarking across organizations, monitoring trends over time, or targeted action to promote long-term workforce sustainability, and national and regional evidence on inequalities in recruitment and career progression among nursing and midwifery staff remains limited.

Applying a clear and reproducible approach to routinely collected workforce data may therefore help strengthen workforce equality monitoring across healthcare settings. We aimed to examine ethnic inequalities in recruitment outcomes and workforce representation across pay bands among nursing and midwifery staff, and to assess whether routinely collected administrative data can generate reproducible indicators for workforce equality monitoring.

## Methods

### Study Design

We carried out a retrospective observational study using routine workforce data from one NHS Board in Scotland. We analyzed two datasets:

- cross-sectional data on staff in post for financial years 2021/22 to 2024/25, and
- pooled administrative data on interviewed candidates and conditional job offers for financial years 2021/22 to 2023/24.

We followed the Strengthening the Reporting of Observational Studies in Epidemiology (STROBE) reporting guidelines (17).

### Setting

In the NHS, pay grades are categorized as “Bands”. Band 5 represents the main entry point to registered practice. Band 6 includes roles requiring specialist skills or leadership responsibilities. Band 7 comprises specialist or managerial positions, while Band 8a and above includes senior roles like nurse consultants or senior managers. Scotland has no national dataset on workforce ethnicity. Analyses therefore relied on Board-level data.

### Data Sources

We extracted staff-in-post data for nursing and midwifery staff from the human resources (HR) systems. We obtained recruitment data from JobTrain, the national NHS Scotland applicant-tracking system, which records interview attendance and conditional job offers. Data on interim recruitment stages were inconsistently recorded; analyses were therefore restricted to interview and conditional offer stages, where data completeness was highest.

We performed secondary analyses of non-identifiable data. In line with UK Health Research Authority guidance, NHS Research Ethics Committee review and NHS Board management approval were not required. We conducted the data extraction and preparation in collaboration with the Board’s Workforce Planning Team, who provided guidance on data quality and completeness.

### Data Categorization

In NHS administrative datasets, the 22 Scottish Census ethnicity categories are aggregated into two broad groups: ‘White’ and ‘Black and Minority Ethnic (BME)’. We therefore restricted the analyses to this pre-existing structure. In place of BME, we used the term non-White, an analytically equivalent but more descriptive label, as the ‘White’ category under PSED encompasses both White British and White minority groups (e.g., Polish, Gypsy/Traveller). In staff-in-post data, we treated records coded as ‘Prefer not to say’ as a distinct analytic category; we did not retain this category in the recruitment dataset due to its low frequency. All analyses were stratified by pay band and ethnic category.

### Variables

#### Outcomes

- Representation across pay bands: quantified using representation quotients (RQs) as a descriptive indicator of relative representation across grades.
- Recruitment success: defined as receipt of a conditional job offer among interviewed candidates.

#### Exposure

- Ethnic category: White vs non-White. In recruitment data, ethnicity was self-reported at application; in staff-in-post data, ethnicity was recorded in the Board’s HR system.

### Data Analysis

After descriptive analyses, we calculated RQs to quantify representation across pay bands, following methods used in prior studies of demographic representation (18,19). The RQ was defined as the proportion of a subgroup within a given pay band divided by its proportion in the total nursing and midwifery workforce. An RQ of 1 indicates equal representation, values above 1 indicate overrepresentation, and values below 1 indicate underrepresentation. Analyses were conducted in Microsoft Excel (Microsoft Corp).

We assessed recruitment disparities by estimating relative risks (RRs) of receiving a conditional job offer at Bands 5, 6, and 7, following the NHS England Workforce Race Equality Standard (WRES) methodology (20). We estimated RRs and 95% confidence intervals (CIs) using 2×2 contingency tables. We excluded staff-in-post records for Band 8a and above due to low counts (non-White n ≤ 5 across all years). We excluded recruitment data for 2024/25 from analysis due to a data quality error identified in the applicant tracking system.

Given the study’s aim of informing policy-relevant workforce equality monitoring, we report unadjusted disparities at key recruitment and workforce decision points. This approach prioritises transparent, descriptive estimates that align with how routinely collected administrative data are typically used in operational monitoring and supports comparability with existing monitoring indicators.

#### Missing data

Ethnicity missingness was 0.6% in recruitment data and ranged from 3.9% to 7.6% in staff-in-post data, with similar levels across years. We conducted the analyses using a complete-case approach, restricted to records with non-missing values for all variables.

## Results

### Staff-in-post Analysis

Between financial years 2021/22 and 2024/25, the proportion of non-White staff at Band 5 increased from 9% to 14% (Table 1). However, representation in higher Bands remained low, with non-White staff comprising 3–4% of the Band 6 workforce and 2% of the Band 7 workforce throughout the period examined. Table 1 summarizes the ethnic distribution of nursing and midwifery staff by White, non-White, and ‘Prefer not to say’ categories across the study period.

**Table 1.**
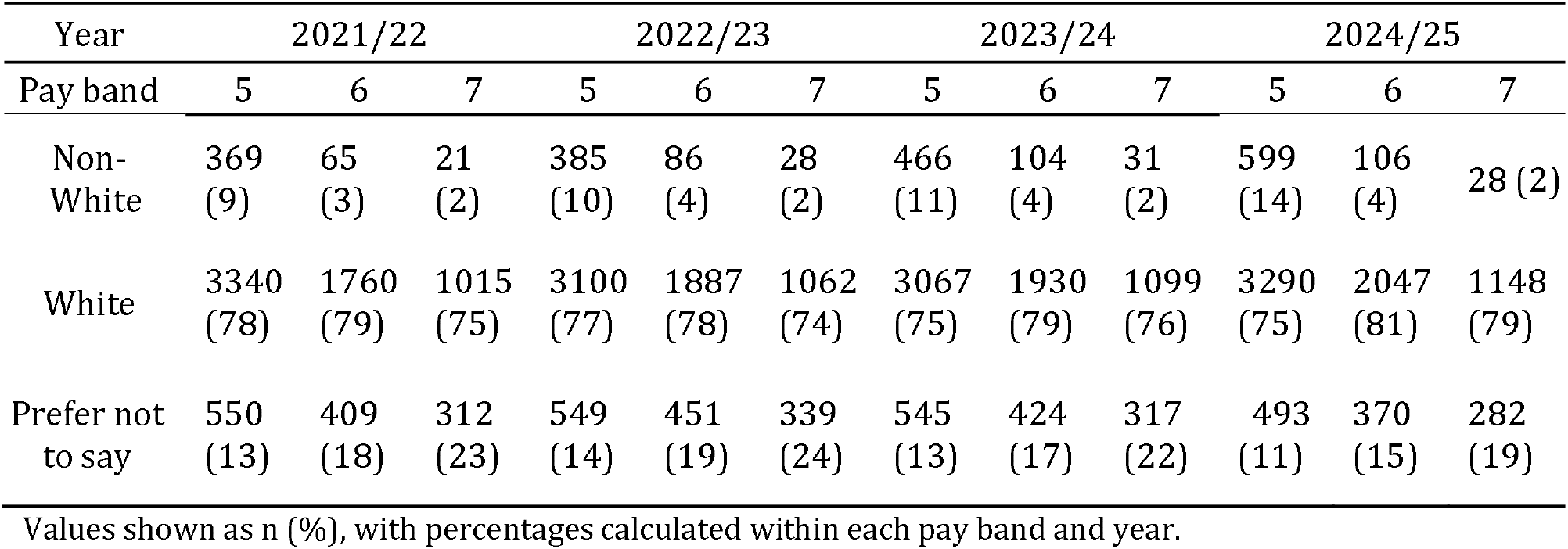
Ethnic distribution of nursing and midwifery staff in post across pay bands.

| Year | 2021/22 |  |  | 2022/23 |  |  | 2023/24 |  |  | 2024/25 |  |  |
| --- | --- | --- | --- | --- | --- | --- | --- | --- | --- | --- | --- | --- |
| Pay band | 5 | 6 | 7 | 5 | 6 | 7 | 5 | 6 | 7 | 5 | 6 | 7 |
| Non-White | 369<br>(9) | 65<br>(3) | 21<br>(2) | 385<br>(10) | 86<br>(4) | 28<br>(2) | 466<br>(11) | 104<br>(4) | 31<br>(2) | 599<br>(14) | 106<br>(4) | 28 (2) |
| White | 3340<br>(78) | 1760<br>(79) | 1015<br>(75) | 3100<br>(77) | 1887<br>(78) | 1062<br>(74) | 3067<br>(75) | 1930<br>(79) | 1099<br>(76) | 3290<br>(75) | 2047<br>(81) | 1148<br>(79) |
| Prefer not to say | 550<br>(13) | 409<br>(18) | 312<br>(23) | 549<br>(14) | 451<br>(19) | 339<br>(24) | 545<br>(13) | 424<br>(17) | 317<br>(22) | 493<br>(11) | 370<br>(15) | 282<br>(19) |

Values shown as n (%), with percentages calculated within each pay band and year.

RQs indicated persistent underrepresentation of non-White staff at Bands 6 and 7, with overrepresentation at Band 5 (Table 2).

**Table 2.**
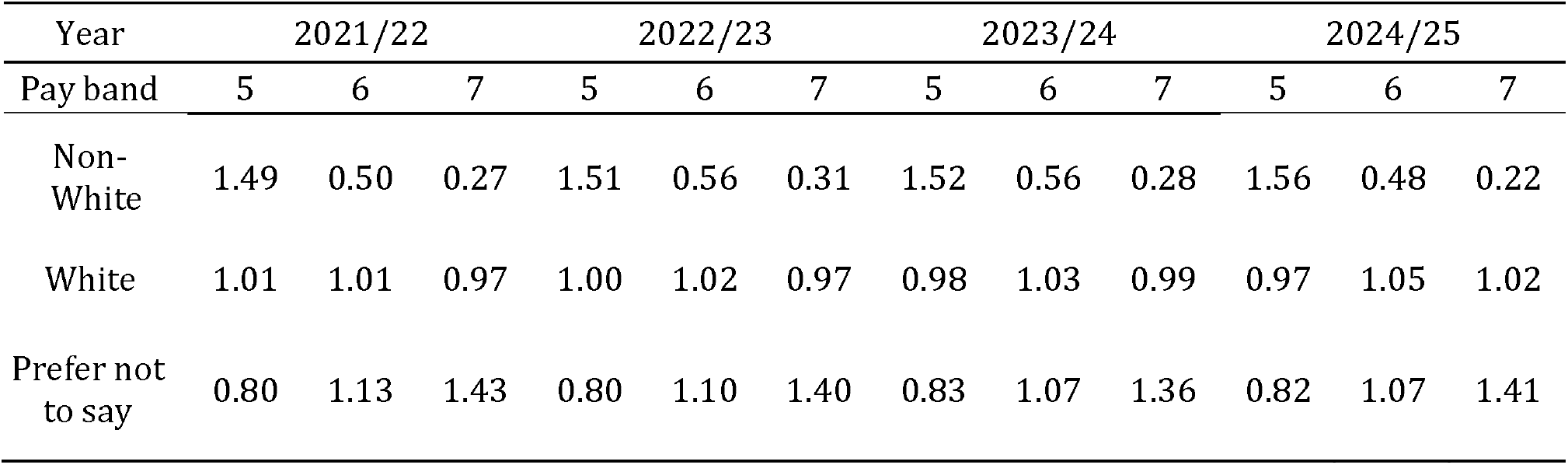
Representation Quotients (RQs) for nursing and midwifery staff in post across pay bands.

**Figure 1.**
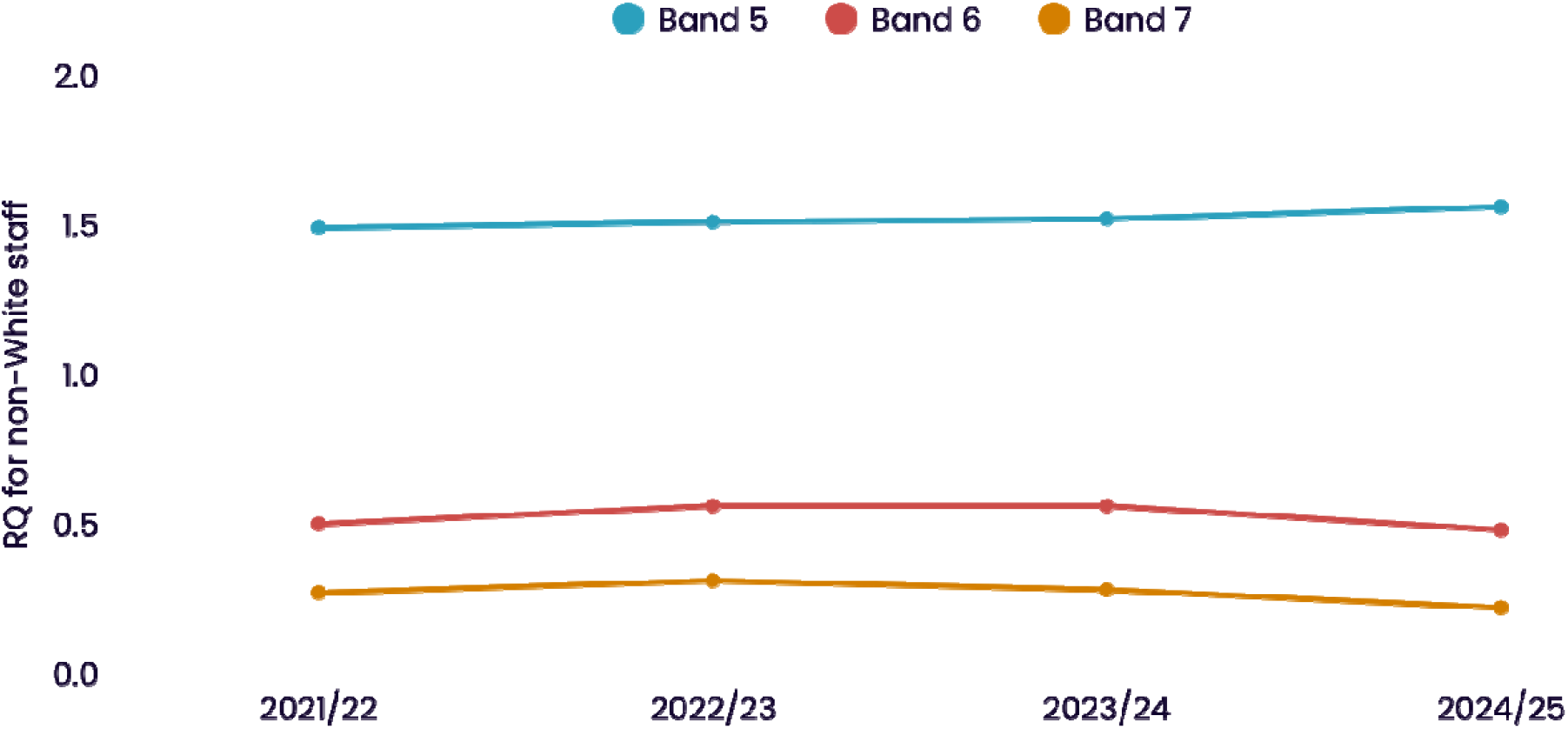
Yearly trends in representation quotients (RQ) for non-White staff across pay bands, 2021/22 to 2024/25. RQ = proportion of non-White staff within a pay band divided by the proportion of non-White staff in the overall nursing and midwifery workforce. Values <1 indicate underrepresentation.

### Recruitment Analysis

Post-interview conditional job offer rates were consistently lower for non-White applicants across the pay bands examined (Table 3). Offer-per-interview rates for Band 5 roles were variable across the study period for both White and non-White applicants, with rates declining markedly in 2023/24. At Band 6, a marked decline was observed only among non-White applicants, from 34% in 2021/22 to 21% in 2023/24. Table 3 summarises interview attendance and conditional job offers for nursing and midwifery posts by ethnic category and pay band.

**Table 3.**
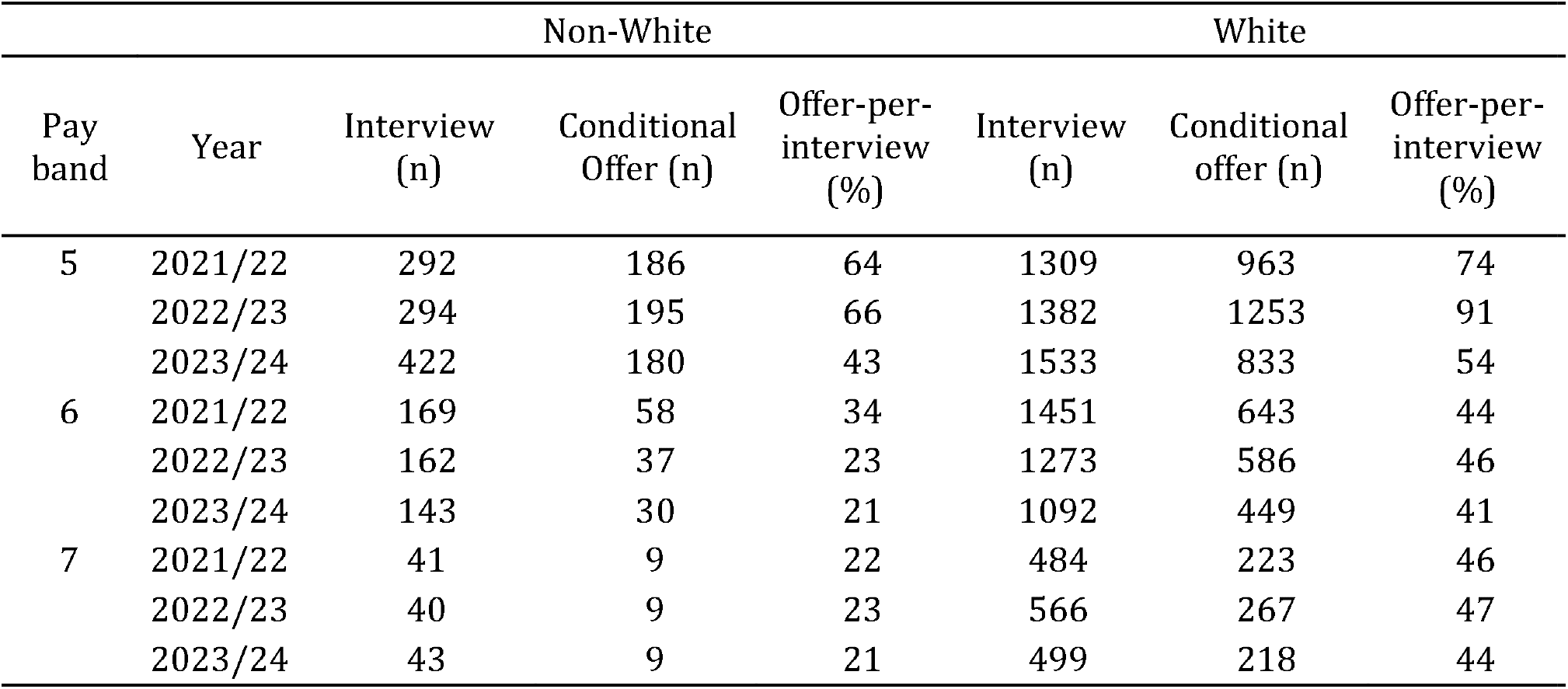
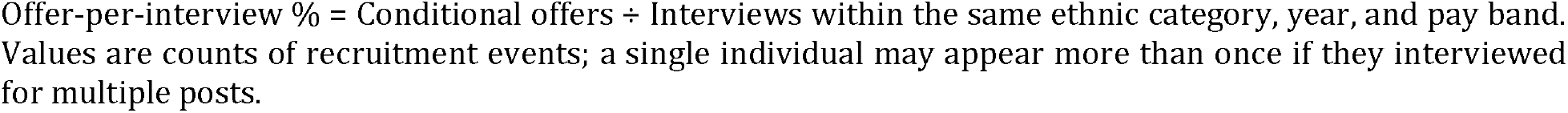
Interview attendance and conditional offers by ethnic category.

| Pay band | Year | Non-White |  |  | White |  |  |
| --- | --- | --- | --- | --- | --- | --- | --- |
|  |  | Interview (n) | Conditional Offer (n) | Offer-per-interview (%) | Interview (n) | Conditional offer (n) | Offer-per-interview (%) |
| 5 | 2021/22 | 292 | 186 | 64 | 1309 | 963 | 74 |
|  | 2022/23 | 294 | 195 | 66 | 1382 | 1253 | 91 |
|  | 2023/24 | 422 | 180 | 43 | 1533 | 833 | 54 |
| 6 | 2021/22 | 169 | 58 | 34 | 1451 | 643 | 44 |
|  | 2022/23 | 162 | 37 | 23 | 1273 | 586 | 46 |
|  | 2023/24 | 143 | 30 | 21 | 1092 | 449 | 41 |
| 7 | 2021/22 | 41 | 9 | 22 | 484 | 223 | 46 |
|  | 2022/23 | 40 | 9 | 23 | 566 | 267 | 47 |
|  | 2023/24 | 43 | 9 | 21 | 499 | 218 | 44 |

Offer-per-interview % = Conditional offers ÷ Interviews within the same ethnic category, year, and pay band. Values are counts of recruitment events; a single individual may appear more than once if they interviewed for multiple posts.

RRs indicated that White candidates were more likely to receive a conditional job offer than non-White candidates (Table 4). Band 6 showed the most pronounced change and worsening disparity, with RR increasing from 1.29 (95% CI 1.04–1.60) in 2021/22 to 1.96 (95% CI 1.41–2.72) in 2023/24. Less pronounced but persistent disparity was also seen at Band 5, the entry point to registered practice.

**Table 4.** Relative Risk (RR) of receiving a conditional offer after interview (White vs non-White)

| Pay band | 2021/22 |  | 2022/23 |  | 2023/24 |  |
| --- | --- | --- | --- | --- | --- | --- |
|  | RR (95% CI) | p | RR (95% CI) | p | RR (95% CI) | p |
| Band 5 | 1.15<br>(1.05–1.27) | <0.001 | 1.37<br>(1.26–1.49) | <0.001 | 1.27<br>(1.13–1.44) | <0.001 |
| Band 6 | 1.29<br>(1.04–1.60) | 0.014 | 2.02<br>(1.51–2.69) | <0.001 | 1.96<br>(1.41–2.72) | <0.001 |
| Band 7 | 2.10<br>(1.17–3.77) | 0.003 | 2.10<br>(1.17–3.76) | 0.003 | 2.09<br>(1.16–3.76) | 0.004 |

RR>1 indicates higher risk for White candidates.

## Discussion

Using routinely collected administrative data from one NHS Board in Scotland, we examined ethnic inequalities in recruitment outcomes and workforce representation among nursing and midwifery staff, and assessed the utility of these data for generating reproducible indicators for workforce equality monitoring. We identified ethnic disparities in recruitment outcomes during 2021/22-2023/24 and persistent underrepresentation of non-White staff in higher pay bands across 2021/22-2024/25. White applicants were more likely to receive a conditional offer following interview across all pay bands examined, with the disparity most pronounced at Band 7 (RRs 2.09–2.10; all p<0.005). In the context of chronic vacancies (21), recruitment inequalities were also evident at Band 5, the standard entry point to registered practice (RRs 1.15–1.37; all p<0.001). Workforce composition analyses showed a corresponding gradient in representation, with non-White staff overrepresented in Band 5 and underrepresented in Bands 6 and 7, with little change over the study period. Taken together, these findings show that routinely collected administrative data can generate clear and reproducible indicators for monitoring ethnic inequalities in recruitment and workforce representation.

To our knowledge, this is the first UK study to combine a representation quotient approach to workforce distribution with analysis of disparities in recruitment outcomes among nursing and midwifery staff. This paired approach strengthens workforce equality monitoring by linking observed disparities at appointment with the corresponding representation gradient in pay band distribution. In 2024/25, non-White staff comprised 14% of Band 5 roles but only 2% of Band 7 roles. Taken together, the recruitment and workforce patterns we observed are consistent with a cumulative disadvantage model, where small inequalities at repeated decision points compound increasingly unequal outcomes across career trajectories (22). This gradient mirrors prior evidence of ethnic disparities in nursing (8,23), and similar gradients reported across other professional groups (24,25).

While the combined recruitment and representation findings demonstrate the potential of routinely collected workforce data for equality monitoring, they also highlight constraints within the underlying data system. We observed inconsistent and incomplete use of status fields within the national applicant tracking system, with some candidate records bypassing ‘shortlisted’ or ‘conditional offer’ stages entirely. As a result, although monitoring frameworks typically compare shortlisting with conditional offer outcomes (20,26), only interview stage records were sufficiently complete for reliable analysis. This precluded examination of potential inequalities that occur at application screening and shortlisting, which have been documented elsewhere (27,28). Such inconsistencies limit the robustness and comparability of equity monitoring and highlight the need for stronger system-level data governance.

Beyond these technical constraints, our findings point to a broader governance gap in routine workforce equality monitoring. Although administrative workforce data are often collected in health systems with mature data infrastructures, they are not always embedded within structured arrangements for the analysis, reporting and benchmarking of equality indicators. Without such arrangements, the potential of routine data to support trend monitoring, organizational accountability and targeted action may remain underused. In Scotland, this challenge is evident in the absence of a comprehensive national framework for workforce equality indicators. This contrasts with developments elsewhere in the UK. In response to longstanding evidence of ethnic inequalities within NHS institutions (9,29), NHS England developed the Workforce Race Equality Standard (WRES), introduced in 2015 as a national reporting and accountability mechanism (14). While implementation has been uneven and the framework has recognised limitations (30), structured monitoring has increased data availability and accountability (26). Wales adopted WRES in 2022 across its health and social care sectors (31). Despite limitations, these approaches illustrate how standardized indicators can support more consistent monitoring and accountability. In the absence of an equivalent framework in Scotland, the routine synthesis and benchmarking of workforce equality data remain fragmented.

### Strengths and Limitations

This study has several strengths. First, it used routinely collected administrative workforce data, demonstrating a pragmatic approach to equality monitoring based on information that health systems already hold. This keeps the analysis close to how workforce inequality monitoring would operate in practice, increasing its policy and operational relevance. Second, it combined analysis of recruitment outcomes with workforce representation across pay bands, allowing inequalities in appointment patterns and workforce distribution to be examined together. Finally, by applying transparent summary measures to data collected over multiple years, the study offers a reproducible approach that could be repeated over time, adapted to other organizational settings, and extended to other protected characteristics where data are sufficiently complete.

Our study has several limitations. First, the analysis was based on routine administrative data from a single NHS organization and should therefore be interpreted as an empirical example rather than a national picture of workforce ethnic inequalities. Second, missing workforce ethnicity data ranged from 4% to 8% across pay bands. Evidence suggests that ethnicity data in administrative systems are frequently not missing at random, with misclassification and non-response more common among minority ethnic groups (32). This pattern would likely attenuate observed associations, meaning that the disparities reported here may be conservative. Finally, the study relied on pooled cross-sectional administrative data. We were therefore unable to track individual career trajectories or make causal inferences. Year-to-year variation in hiring patterns may also influence band-level representation. These limitations do not diminish the value of routine data for workforce equality monitoring, but highlight the importance of improving data completeness, quality and longitudinal linkage.

### Recommendations for Further Research

Further research should examine the applicability of this approach across multiple organizations, health systems and national contexts using routine administrative data. Such work would help assess whether similar indicators can be generated consistently in different settings, and identify the minimum standards needed to support wider use of routine workforce equality monitoring in settings without an equivalent mechanism. Research should also examine whether additional indicators can be derived reliably from routine workforce systems, and whether these are feasible and useful for monitoring inequalities in practice. Longitudinal linkage could further strengthen monitoring of inequalities over time.

### Implications for Policy and Practice

The findings have implications for both workforce policy and organizational practice. They suggest that routinely collected administrative data provide a practical basis for monitoring ethnic inequalities in recruitment and workforce representation, despite imperfections in the data systems. Health organizations should therefore move beyond descriptive reporting of workforce equality data towards more systematic use of standardized indicators, with relevance not only to equity and accountability but also to workforce sustainability and retention. In Scotland, where substantial administrative workforce data are already collected, the priority is less the creation of entirely new data sources than the development of clearer frameworks for routine analysis, reporting and benchmarking. More broadly, health systems seeking to strengthen workforce equality monitoring should prioritise improved ethnicity data quality, more consistent recording of recruitment stages, and governance arrangements that support repeated, comparable analysis over time.

## Conclusion

Using routinely collected administrative data from one NHS organization in Scotland, we demonstrate a clear and reproducible approach to monitoring ethnic inequalities in recruitment and workforce representation among nursing and midwifery staff. By combining recruitment outcome data with representation quotients, it illustrates how complementary indicators can be derived from existing workforce systems and applied systematically over time. Using this approach, we identified ethnic inequalities in recruitment outcomes and persistent underrepresentation of non-White staff in more senior pay bands. Embedding such standardized analyses within routine workforce monitoring could strengthen benchmarking, accountability and targeted action across healthcare settings. Given evidence linking workforce inclusion to staff experience, retention and patient care (7,33,34), making better use of existing workforce data represents a pragmatic step towards more equitable and sustainable health services.

## Supporting information

STROBE checklist

## Data Availability

All data produced in the present study are available upon reasonable request to the authors.

